# Does Clinical Nihilism affect the diagnostic approach for patients with undiagnosed adult coeliac disease? A UK Multicentre study

**DOI:** 10.1101/2020.08.28.20181750

**Authors:** RJ Blanshard, G Naylor, MA Taylor, HA Penny, PD Mooney, DS Sanders

**Author notes:** **Corresponding Author:** Professor David Sanders, Consultant Gastroenterologist, Royal Hallamshire Hospital, Glossop Road, Sheffield, United Kingdom.

## Abstract

**Background:** Adult coeliac disease (CD) has delays in diagnosis but the reasons for this have not been explored.

**Methods:** Group 1) Time from primary care presentation to diagnostic endoscopy was prospectively quantified in 151 adult patients with a positive endomysial antibody test and compared with the diagnostic pathway of 92 adult patients with suspected inflammatory bowel disease (IBD). Group 2) Across 4 hospitals over a 3-month period, duodenal biopsy reports for suspected CD were reviewed (n=1423). Group 3) 50 gastroenterologists completed questionnaires concerning their viewpoints on CD.

**Results:** Group 1) Suspected coeliac patients waited significantly longer for diagnostic endoscopy following referral (48.5 [28-89] days) than suspected IBD patients (34.5 [18-70] days; p=0.003). Group 2) Of the 1423 patients that underwent diagnostic endoscopy for possible CD, 40.0% met the guidelines to take at least 4 biopsies. Diagnosis of CD was more likely if these guidelines were followed (10.1% vs 4.6% p<0.0001). 12.4% of newly diagnosed CD patients had at least 1 non-diagnostic gastroscopy in the 5 years prior to diagnosis. Group 3) 32.0% (16) of gastroenterologists failed to identify that CD has greater prevalence in adults than IBD. Moreover, 36.0% (18) of gastroenterologists felt that doctors were not required for the management of CD.

**Conclusion:** Prolonged waiting times for endoscopy and inadequacies in biopsy technique suggest clinical inertia towards CD. This is exemplified by the nihilistic approach to the condition demonstrated in our qualitative data. This is the first study to demonstrate clinical inertia towards CD.

**Main messages:**

- CD patients face greater delays at all wait intervals from referral to diagnostic endoscopy compared to inflammatory bowel disease (IBD) patients.
- The majority of endoscopists do not follow guidelines for diagnostic endoscopy for CD. This reduced diagnosis rates by over 50%.
- Questionnaire findings presented a nihilistic attitude towards CD alone and in comparison to IBD.
- This is the first study to fully represent how clinical inertia towards CD directly leads to increased diagnostic delay and likely missed diagnoses.

**What is already known on the subject:**

- Diagnostic delay is a key issue facing modern management of coeliac disease (CD) whilst only 1 in 4 cases are estimated to be diagnosed.
- Delayed diagnosis of CD is associated with an increased risk of complications from the disease and a worse quality of life.

## Introduction

Global meta-analysis of screening studies for coeliac disease (CD) has shown a variable prevalence of around 1%,^1-4^ but the vast majority of these patients remain undiagnosed.^1,5,6^ Comparison of screening studies with point prevalence data has demonstrated that only an estimated 1 in 4 cases of CD are diagnosed in the UK,^5^ representing a significant undiagnosed burden. However, in other countries such as the US, studies have reported this undiagnosed burden to be far greater at 80%-90%.^7,8^

Compounded with this undiagnosed burden, diagnostic delay is a key and widely reported issue facing modern management of CD.^9-12^ Delays in diagnosis can present in both primary and secondary care and international literature reports a mean delay range of 9.7-12.8 years.^13-16^

Of particular concern, these delays persist despite improvements in factors such as serological test availability, ease of access to endoscopy and increasing public awareness of the condition.^5^ These long delays are especially worrying in context of the substantial improvement to quality of life associated with diagnosis.^9,16^ Despite this burden of missed diagnoses and diagnostic delay, the incidence of CD continues to rise, with a fourfold increase in incidence seen in the UK between 1990-2011,^5^ overall representing an increasing public health problem.

Delayed diagnosis of CD is correlated with increased risk of complications such as osteoporosis, peripheral neuropathy, microcytic (iron deficiency) and macrocytic (folate deficiency) anaemia, lymphoma (enteropathy associated T-cell lymphoma (EATL) and other non-hodgkin lymphomas), hyposplenism, micronutrient deficiencies and increased rates of anxiety and depression.^11,17^ Importantly, earlier diagnosis is associated with lower standardised mortality ratios,^18,19^ whilst undiagnosed CD is associated with a worse quality of life which improves substantially upon diagnosis.^9,16^ Prompt management with a strict gluten free diet (GFD) leads to a rapid reduction in symptoms and reduced risk of malignancy (specifically gastrointestinal carcinoma or lymphoma).^11,17^

Previous studies investigating the cause of such delay and undiagnosed burden have hypothesized reasons such as patient related factors,^10^ to inability by clinicians to recognise the extraintestinal symptoms as indications for serological testing.^8,20^ More recently, the focus on causes of delay has centred around clinician factors such as clinical inertia towards the condition by primary care physicians and inadequacies in biopsy technique in diagnosis of the condition. A study of primary care physicians in the USA found only 60% would perform serological testing for a young caucasian man with unexplained IDA whilst 80% said they would start a serologically positive patient on a GFD prior to endoscopy. This suggests a failure to recognise the pervasiveness of CD and a lack of clinical knowledge in how to properly approach diagnosing the condition, suggesting a level of clinical inertia towards it.^21^ Meanwhile, studies examining previous endoscopies in CD patients demonstrated a failure to complete biopsies or take an adequate number of biopsy samples despite presence of symptoms suggestive of CD prior to diagnosis, resulting in delayed diagnoses.^22,23^

In attempts to deal with these issues of delayed and missed diagnoses, guidelines for the management of CD are constantly updated to account for the evolving knowledge base on this disease. Current National Institute for Health and Care Excellence (NICE) guidelines recommend that patients referred with suspected CD have a duodenal biopsy within 6 weeks of referral and that the patient is strongly encouraged to eat gluten in more than one meal a day for at least 6 weeks before the procedure.^24^ This may prove problematic for patients who have self-diagnosed themselves with CD and are already abstaining from gluten. British Society of Gastroenterology (BSG) guidelines indicate that during diagnostic endoscopy at least 4 biopsy specimens should be taken, including a duodenal bulb biopsy.^25^ This is in order to maximise diagnostic yield, as taking at least 4 biopsy specimens is shown to more than double the diagnostic rate in comparison to those undergoing less than 4 biopsies.^23,26^ Whether these guidelines are adhered to in clinical practice remains an important question.

There is a lack of contemporary UK data assessing if there is still a delay in diagnosis. Moreover, reasons for delay are yet to be comprehensively explored. To refine the diagnostic pathway for CD, these factors must be characterised. The aim of this UK multicentre study was to assess the degree of delay present in the diagnostic referral pathway for CD as well as determine concordance with biopsy guidelines. Additionally, clinician attitudes towards CD were explored in an attempt to characterise the factors influencing delay.

## Materials and Methods

### Group 1: Primary care presentation to biopsy completion and case complexity

All patients who registered a positive endomysial antibody (EMA) test over an 18 month period in the South Yorkshire area were assessed for inclusion eligibility. The following data on each patient was collected: Hospital ID, Date of Birth, Gender, date of initial EMA positive blood test in primary care, date of referral to secondary/tertiary care, department referred to (gastroenterology, endoscopy, other), dates of interval appointments, date of endoscopy and Marsh Grade of duodenal biopsy specimens. For each patient, archived blood tests were examined to identify their first EMA positive result, which was recorded as the initial positive EMA result to be used in exclusion criteria. Patients were excluded if they were aged under 16, had a previously known diagnosis of CD, if the initial positive EMA result was requested by the gastroenterology outpatients department or other speciality (other than primary care) or if the patient never received a duodenal biopsy.

For the control group, all patients with a histological diagnosis of inflammatory bowel disease (IBD) following colonoscopy or flexible sigmoidoscopy were examined. The same data as with suspected CD patients regarding dates of referral, appointments and endoscopy was collected and the same exclusion criteria applied. This selection criteria produced a cohort of 151 suspected CD patients and 92 IBD patients.

An additional observational study was also completed to compare patients attending a specialist CD clinic in a central teaching hospital (n=102) over a six month period against two control groups: patients attending an established specialist IBD service in a central teaching hospital (n=99), and patients with a diagnosis or suspected diagnosis of CD (n=36) attending a general gastroenterology clinic at a district general hospital. This additional study analysed clinic appointment complexity. Data regarding patient presenting symptoms, investigations (bloods, imaging, endoscopies, other), medications, referrals completed and planned follow up periods was collected from clinic appointments.

### Group 2: Adherence to biopsy guidelines in detection of CD

Endoscopy and histology reports for all patients who had a duodenal biopsy for suspected CD in a 3 month period in 4 UK hospitals were retrospectively reviewed. Indications for biopsy, number of specimens received by histopathology, biopsy technique (single bite versus double bite), job roles of the endoscopist (physician, surgeon or nurse endoscopist) and final diagnosis were recorded for this cohort of 1423 patients. Findings of villous atrophy were required for diagnosis of CD. Of those subsequently diagnosed with CD, patient records were checked for any previous non-diagnostic gastroscopies in the 5 years prior to diagnosis. Patients were excluded if they had known CD.

### Group 3: Gastroenterology clinicians’ perspective

A questionnaire aimed at assessing clinician attitudes towards CD was formed through use of a focus group consisting of a range of clinicians specialised in gastroenterology. Discrete choice experiments (DCEs) were used to establish clinician preferences using multiple options and ranking for interventions and service provision in the comparison of CD with Crohn’s disease and ulcerative colitis (UC). Direct questioning (Yes or No) was used to determine if the opinions of the focus group matched those gastroenterology clinicians.

A proforma (see supplementary) was completed through discussions with 50 gastroenterology registrars and consultants from across the UK. Staff grades and specialist interests of clinicians completing the survey were also recorded.

## Results

### Group 1: Primary care presentation to biopsy completion and case complexity Time from referral to endoscopy

Time from referral to endoscopy was 48.5 [28-89] days for CD patients from both centres combined. This was significantly longer than the wait for suspected IBD patients (34.5 [18-70] days; p=0.003).

#### Biopsy grade

Longer delays to diagnosis correlated with a significantly decreased Marsh Grade of duodenal biopsies. Of those seen within 4 weeks, 15.5% had normal-borderline (Marsh Grade 0-3a) histology results, compared to 44.9% seen after 4 weeks (p=0.017).

#### Case complexity

Analysis of case complexity data (mean number of reports of abdominal pain, diarrhoea, stool urgency, bloating, bleeding per rectum (PR)/in stool, nausea/vomiting, headache, weight loss and fatigue) showed there was no significant difference in the number of times each presenting complaint was discussed and in the mean number of presenting complaints reported per patient between the CD group and the general clinic control group.

When comparing the two specialist clinics the following investigations were requested significantly more in the CD clinic than in the IBD clinic: Blood tests, genetic testing, gastroscopy and DXA scans (Table 1). When using the Bonferroni correction to adjust for multiple analysis the p-values remained less than 0.05 and therefore remained significant. When using the Bonferroni correction, no investigations were requested for more patients in the IBD group compared to the CD group.

**Table 1:** Investigations at clinic appointments.

|  | Coeliac | IBD | P-value | Adjusted P-value | General | P-value | Adjusted P-value |
| --- | --- | --- | --- | --- | --- | --- | --- |
| **Bloods** | 83 (81.4%) | 43 (43.4%) | **<0.001** | **<0.001** | 26 (72.2%) | 0.247 | - |
| **Genetic Testing** | 13 (12.7%) | 0 (0%) | **<0.001\*** | **0.002** | 3 (8.3%) | 0.562* | - |
| **Gastroscopy** | 21 (20.6%) | 1 (1.0%) | **<0.001** | **<0.001** | 6 (16.7%) | 0.610 | - |
| **Colonoscopy** | 7 (6.9%) | 16 (16.2%) | **0.038** | 0.38 | 1 (2.8%) | 0.680* | - |
| **Flexible Sigmoidoscopy** | 1 (1.0%) | 3 (3.0%) | 0.364* | - | 0 (0%) | 1.000* | - |
| **Capsule Endoscopy** | 2 (2.0%) | 5 (5.1%) | 0.274* | - | 0 (0%) | 1.000* | - |
| **Imaging** | 4 (4.0%) | 10 (10.1%) | 0.085 | - | 1 (2.8%) | 1.000* | - |
| **SeHCAT Test** | 2 (2.0%) | 2 (2.0%) | 1.000* | - | 0 (0%) | 1.000* | - |
| **DXA Scan** | 15 (14.7%) | 0 (0%) | **<0.001** | **<0.001** | 5 (13.9%) | 0.905 | - |
| **Breath Tests** | 2 (2.0%) | 3 (3.0%) | 0.678* | - | 0 (0%) | 1.000* | - |
| **Mean number of investigations** | 2.19 (+/- 1.41) | 1.93 (+/- 1.35) | **<0.001** | - | 1.44 (+/- 1.23) | 0.054 | - |

\*= One or more of the cells in 2x2 table have an expected count <5, Fisher's Exact Test is used to calculate p-values.

Table 2 shows that when using the Bonferroni correction there was no significant difference between the amount of times each medication type was prescribed between the CD and general clinic group. When comparing the CD and IBD groups, a significantly greater number of patients were prescribed immunosuppressive medication, disease-modifying antirheumatic drugs (DMARDs) and anti-inflammatory medication.

**Table 2:** Prescribed medications at clinic appointments.

|  | Coeliac | IBD | P-value | Adjusted P-value | General | P-value | Adjusted P-value |
| --- | --- | --- | --- | --- | --- | --- | --- |
| **Steroid** | 16<br>(15.8%) | 21<br>(21.2%) | 0.312 | - | 0 (0%) | **0.012\*** | 0.084 |
| **Immuno-suppression** | 16<br>(15.8%) | 38<br>(38.4%) | **<0.001** | **0.002** | 1 (2.8%) | **0.043\*** | 0.301 |
| **DMARD** | 1 (1.0%) | 21<br>(21.2%) | **<0.001** | **<0.001** | 0 (0%) | 1.00* | - |
| **Anti-Inflammatory** | 0 (0%) | 19<br>(19.2%) | **<0.001** | **<0.001** | 0 (0%) | 1.00* | - |
| **Anti-Spasmodic** | 5 (5.0%) | 1 (1.0%) | 0.212* | - | 0 (0%) | 0.326 | - |
| **Sequestrant** | 2 (2.0%) | 2 (3.0%) | 1.00* | - | 0 (0%) | 1.00* | - |
| **Other** | 11<br>(10.9%) | 11<br>(11.1%) | 0.820* | - | 1 (2.8%) | 0.288 | - |

\*= One or more of the cells in 2x2 table have an expected count <5, Fisher's Exact Test is used to calculate p-values.

### Group 2: Adherence to biopsy guidelines in detection of CD

Of the 1423 patients that underwent duodenal biopsy, 97 (6.8%) of these were subsequently diagnosed with CD. Regarding biopsy guidelines, 40.0% the total number of patients who underwent diagnostic endoscopy had at least 4 biopsies taken. The median number of biopsies taken per patient was 3. If guidelines to take at least 4 biopsy samples were followed, diagnosis of CD was more likely than if 3 or less biopsy samples were taken (10.1% vs 4.6% p<0.0001). Whilst the median number of biopsies was greater in patients diagnosed with CD (4 vs 3 p<0.0001).

Of the patients that received a CD diagnosis following biopsy, 12.4% had received at least 1 non-diagnostic gastroscopy (describing a gastroscopy where no biopsy samples were taken) in the 5 years prior to diagnosis. When assessing endoscopist job roles, gastroenterologists and nurse endoscopists were significantly more likely than surgeons to follow guidelines (41.5% vs 51.2% vs 18.2% p<0.0001) and therefore took at greater number of biopsies (3 vs 4 vs 2, p<0.0001). Thus, gastroenterologists and nurse endoscopists made a diagnosis of CD in more cases than surgeons (7.1% vs 6.7% vs 3.0%, p=0.10). The use of single bite biopsy technique compared to double bite resulted in an increase of 3 to 4 biopsies (p=0.02) taken from the second part of the duodenum (D2).

### Group 3: Gastroenterology clinicians’ perspective

Of those completing the questionnaire, 64.0% (32) were registrar grade (trainee) and 36.0% (18) were consultants.

Questionnaire results revealed that 32.0% (16) of gastroenterologists failed to identify that CD has greater prevalence in adults than IBD. 36.0% (18) of gastroenterologists felt that doctors were not required for the management of CD whilst 16.0% (8) felt that a diagnosis of CD does not significantly impact patient quality of life.

Additionally, 40.8% (20) said that management of CD is not academically challenging. 88.0% (44) believed CD was less difficult to manage than IBD whilst 82.0% (42) thought CD was less significant than IBD in terms of resources needed to diagnose and treat. 77.1% (37) thought CD had less of an impact on quality of life than IBD.

## Discussion

This contemporary multicentre UK study demonstrates prolonged waiting times for endoscopy and inadequacies in biopsy technique for adult patients with suspected CD. This objectively suggests clinical inertia towards CD. This is further supported by the nihilistic approach to the disease demonstrated in our qualitative data. Enhanced knowledge of the clinical significance of CD and improved adherence to biopsy guidelines will result in patients receiving more prompt diagnoses and therefore better health outcomes.

Endoscopy waiting times were not in keeping with NICE guidelines of 6 weeks.^24^ The significant negative correlation between delay till endoscopy and Marsh Grade of biopsy suggests that with greater wait time patients are more likely to self impose a GFD, likely in attempt to achieve the improved quality of life from switching to a GFD,^9,16,27^ increasing the chance of a missed diagnosis. This means it is more likely for these borderline patients to need a repeat biopsy, which subjects these patients to persisting symptoms as they wait to go onto a GFD and causes distress due to clinical uncertainty and having to go through the endoscopy process.

The fact that in only 40.0% of endoscopies 4 biopsy samples were taken clearly represents how in the majority of duodenal biopsies BSG guidelines are not followed.^25^ The detriment from this is clearly shown through the greater than doubled diagnosis rate (10.1% for 4 or more biopsies vs 4.6% for 3 or less) seen in endoscopies where the guidelines were followed. This is a clear example of the impact of diagnostic inertia towards CD on missed or delayed diagnoses and is backed by a similar low rate of guideline adherence (35%) in a USA based biopsy study.^23^ Patients with missed CD will go on to experience more harm through possible repeat endoscopies and more time spent following a gluten containing diet. To build on this, 12.4% of the patients diagnosed with CD had received a previous non-diagnostic (here meaning that no biopsy samples were taken) gastroscopy in the 5 years before their diagnosis.

A strong majority of 88.0% rated IBD as having greater complexity than CD and 82.0% believed CD was less significant than IBD in terms of resources needed to diagnose and treat. Furthermore, 36.0% of clinicians felt that doctors are not required for management of CD and 40.8% believed management of CD was not academically challenging. These findings suggest significant proportions of gastroenterology clinicians believe CD should be treated with less urgency than IBD and that it is not a condition for which medical expertise is required, further building a picture of medical nihilism towards the condition.

A systematic review estimated prevalence of 0.005%-0.5% for UC and 0.0006%-0.32% for Crohn’s disease in Europe.^28^ However, 32.0% of clinicians incorrectly identified IBD as having greater prevalence than CD. A previous study compared Crohn’s disease, UC, CD patients and healthy controls using the Short-Form 36 (SF-36)-Item Health Survey and the Hospital Anxiety and Depression Scale (HADS) to determine quality of life scores. The total study population was 1031, with over 200 patients included for each condition. Although Crohn’s disease patients reported the worst scores for general health, UC patients reported better general health than CD patients.^29^ The influence of diet on social interaction is well characterised and it has been shown patients with CD feel negatively controlled by their dietary restrictions, causing a significant impact on their social relationships^30^ in addition to their physical symptoms. The economic impact of purchasing gluten free products, which can be an average of 4.1 times more expensive and much less available than their gluten containing counterparts is also well recorded, providing an example of the continuing impact of the condition even following effective treatment.^31^

Despite this, 77.1% of gastroenterologists questioned believed CD had less of an impact on patient quality of life than both Crohn’s disease and UC. Moreover, 16.0% of gastroenterologists believed CD caused no significant impact on quality of life. This is a significant proportion considering the specialism of those surveyed and the presence of numerous studies characterising the effect of CD on patient quality of life.^9,16,27^ These findings clearly exhibit medical nihilism towards the condition, showing medical professionals do not appreciate the impact of the condition both alone and in comparison to similar conditions.

This is the first study to objectively demonstrate how medical nihilism translates to clinical inertia which itself causes diagnostic delay in the management of CD. Improved knowledge of the clinical significance of CD may lead to increased referral urgency. Greater adherence to biopsy guidelines will produce higher diagnosis rates. These changes will contribute to reducing the vast undiagnosed burden of CD by producing more prompt diagnoses, resulting in better health outcomes.

## Data Availability

All necessary data can be made available.

## Acknowledgements

None.

